# Orbito-frontal cortex functional connectivity mediates the relationship between fetal growth and childhood impulsivity

**DOI:** 10.1101/2025.09.16.25335932

**Authors:** João Paulo Maires Hoppe, Olivia Ruge, Roberta Dalle Molle, Guillaume Elgbeili, Qizhou Xia, Patricia Pelufo Silveira

**Author notes:** <u>Corresponding author:</u> Patricia Pelufo Silveira, MD, PhD, Department of Psychiatry, Faculty of Medicine, McGill University, Douglas Research Centre, 6875 Boulevard LaSalle, Montreal, QC, H4H 1R3, Canada, (ext.2776).

## Abstract

The prenatal period is critical for a healthy development, and exposure to adversity during it may provoke alterations of several biological tissues and systems, resulting in health outcomes that may take place into childhood and adulthood. The orbito-frontal cortex (OFC), central in cognitive processes, is sensitive to negative environmental effects in the intrauterine environment. We investigated the association between OFC function and decision-making behavior in response to a poor-quality prenatal environment. We evaluated a subsample of the MAVAN longitudinal Canadian birth cohort gathering data on anthropometric measurements at birth, and resting-state functional MRI (rsfMRI) and decision-making (using the Information Sampling Task from the CANTAB battery) measured later in life. We performed a mediation analysis to investigate the direct and indirect effect of being born small for gestational age (SGA) on the Information Sampling Task performance, through OFC-related functional connectivity. Being born SGA is associated with decreased functional connectivity between the left hemisphere OFC and the middle frontal gyrus (OFC–MFG). Additionally, increased OFC–MFG connectivity is linked to better IST performance. Thus, SGA individuals have an altered OFC– MFG functional connectivity, which impacts on their performance on a decision-making task. Lower OFC–MFG functional connectivity and impulsive decision-making were associated to the SGA condition, reflecting a poor-quality prenatal environment. These findings highlight the importance of the prenatal period for a healthy development and suggest that neuroimaging focusing on the affected areas may identify individuals at higher risk of developing psychopathologies, and direct for proper interventions.

## INTRODUCTION

The prenatal period is a critical window to the establishment of a healthy development during childhood, characterized by high plasticity and sensitivity to environmental influences. Intrauterine exposure to adversity may induce alterations in the development of biological systems, with significant consequences for health outcomes during childhood and even well into adulthood (1, 2). It is estimated that one in every four newborns worldwide experiences significant intrauterine adversities that impact their growth and development (3). Prenatal adversity may arise from several factors, such as poor maternal nutrition, chronic diseases (e.g., hypertension, type 2 diabetes, inflammation), and substance abuse during pregnancy, resulting in fetal growth restriction (3). Individuals that are born with a lower birthweight than the expected for their sex and gestational age are classified as small for gestational age (SGA). Thus, being born SGA is a biomarker for a poor-quality prenatal environment (3, 4). Being born SGA is a risk factor for increased predisposition for a plethora of disorders, including cognitive impairments (5, 6), obesogenic behaviors (7, 8), and metabolic and psychiatric disorders (9-13).

The orbito-frontal cortex (OFC) is a brain mesocortical area located in the ventral surface of the frontal lobe. The OFC is a major brain hub, with a wide range of connections including the anterior cingulate cortex (14), ventral striatum (15), hippocampal memory system (16), amygdala, and the brain stem (17, 18). As such, it plays a key role in various cognitive processes, including emotional regulation, value-encoding, and reward-based decision making (19, 20). Disrupted OFC function is linked to increased vulnerability for mental health problems, such as depression, substance abuse, and anxiety disorders (21, 22). The development of the OFC begins early in humans, starting as early as 17 weeks of gestation. OFC synapse formation begins during the prenatal period, peaks postnatally, and is then followed by synaptic pruning (23), and OFC myelination begins during childhood and extends well into young adulthood. Therefore, the OFC is one of the last brain regions to fully develop (24), and this prolonged maturation process makes this brain region sensitive to negative environmental effects for longer during development, particularly from the prenatal period until the first year of life (25, 26).

Studies have documented a range of detrimental effects of early-life adversity on the whole brain anatomy and function (27-34), but also showing alterations in the frontal areas, particularly the OFC. A reduced OFC volume was observed both in children who suffered physical abuse (35), and decreased OFC cortical thickness, as well as increased depressive symptoms, was detected in young adults following early life stress exposure (36). A study investigating fetal growth restriction and resting-state functional connectivity (rsFC) between regions linked to reward, self-control, and value-encoding has demonstrated that being born SGA is associated with altered rsFC between the OFC and the dorsolateral prefrontal cortex both in children and adolescents (7). The mid and long-term effects observed in these studies suggests a lingering influence of early life adversity on the OFC development and behavior. In summary, there seems to be a tripartite association between early life adversity, behavioral alterations, and disrupted OFC function (20), but the nature of this association remains unclear.

Here, we aimed at identifying whether and how neurofunctional and behavioral phenotypes are linked in response to prenatal stress. We hypothesized that this association has an ontological framework, where being born SGA alters OFC function, which in turn leads to impaired decision-making processes. For that, we employed a mediation analysis design, using a population-normed birth weight classification, rsFC of OFC-based functional elements, and a behavioral measurement of decision-making processes (Information Sampling Task) in a Canadian children cohort.

## METHODS

### Study sample

We used mother-infant dyads data from the Maternal Adversity, Vulnerability and Neurodevelopment (MAVAN), a Canadian prospective birth cohort (37). Most women were recruited at the time of routine ultrasound examinations, 16-20 weeks. Exclusion criteria were severe chronic illness requiring ongoing treatment, placenta previa, a history of incompetent cervix, impending delivery, or had offspring born with <37 gestational weeks or with a major anomaly. MAVAN recruited 630 mother-child pairs at two sites, Montreal (Quebec) and Hamilton (Ontario), with longitudinal assessments from the prenatal period to 10-12 years of age. The MAVAN Project was approved by the institutional review boards at hospitals and university affiliates: McGill University, l’Université de Montreal, Royal Victoria Hospital, Jewish General Hospital, Centre Hospitalier de l’Université de Montreal, Hôpital Maisonneuve-Rosemont, St. Joseph’s Hospital, and McMaster University. Informed consent was obtained from the parents/guardians of the participants. Research Ethics Board approval was received from the Douglas Mental Health University Institute (IUSMD-03-45, IUSDM-13-09). In the current study, only children from Montreal were included.

### Birthweight classification

We collected gestational age and birthweight data from each child’s health records to calculate the birth weight ratio (BWR). The BWR is the ratio between the observed birthweight and the sex-specific median birthweight for that gestational age, based on a Canadian population norm (38). Participants were classified as SGA when their BWR was <0.85 (39). Values between 0.85 and 1.15 resulted in an appropriate for gestational age (AGA) classification, whereas BWR >1.15 was classified as large for gestational age (LGA).

### Neuroimaging

Data were acquired using a 3T Trio Siemens scanner (age = 9.07 ± 1.46 years). High resolution T1-weighted images for the whole cerebrum were obtained in an approximately five-minute session (1 mm isotropic 3D MPRAGE, sagittal acquisition, 256 x 256 mm grid, TR = 2300 ms, TE = 4 ms, FA = 9 degrees). Resting-state data were obtained on a six-minute run of blood oxygenation level-dependent (BOLD) signal, at rest, using a gradient echo-planar sequence (TR = 2000 ms, TE = 30 ms, flip angle = 90 degrees) with 3 x 3 mm in-plane resolution with 33 slices of 4 mm on a 64 x 64 matrix. Participants were instructed to stay with their eyes open and fixating on a “+” at the center of the screen, and to relax and avoid thinking about anything. Images were pre-processed using Statistical Parameter Mapping (SPM v.12, University College London, UK; http://www.fil.ion.ucl.ac.uk/spm) in conjunction with the default processing pipeline using CONN Functional Connectivity Toolbox (CONN toolbox; www.nitrc.org/projects/conn). Images were converted from DICOM to NIFTI-1 for processing. To compensate for different temporal offsets between slice acquisition, interpolation (slice-timing correction) was applied to the functional images (40), followed by movement-related artifacts correction. This process (https://web.conn-toolbox.fmri-methods/preprocessing-pipeline) involves identification of outliers via calculation of a framewise displacement of each timepoint within a 140 x 180 x 115 mm bounding box. If displacement exceeds 0.9 mm, it is flagged as a potential outlier. Also, the global BOLD signal change is computed at each timepoint, and if the average BOLD signal changes is greater than 5 s.d., it is also flagged as an outlier. Each image was transformed via a 6-parameter rigid-body transformation, and then an autoregressive moving average model was applied to correct for changes in head positions (41). Then, the corrected images were co-registered to the individual raw T1 anatomical images (Montreal Neurological Institute; MNI152). High-resolution anatomical images were segmented into grey matter, white matter, and cerebrospinal fluid; smoothed gray matter images were then transformed into a 6-parameter three-dimensional quadratic deformation (42, 43). Functional data were smoothed using 8 x 8 x 8 mm Full Width at Half Maximum (FWHM) Gaussian kernel for statistical analysis (44). Resting-state images were processed using CONN toolbox (45). Functional imaging signal was filtered using a temporal band-pass filter of 0.01-0.08 Hz. Residual motion correction parameters were used as regressor in the model according to Behzadi et al. (46) method to mitigate motion-related artifacts. The aCompCor method, part of the CONN pipeline, identifies confounding effects with noise from cerebral white matter and cerebrospinal components as well as head motion. Factors that are identified as confounds were then removed from volumes using Ordinary Least Square regression. Regions of Interest (ROIs) were extracted from Desikan et al. (47) and from the Harvard-Oxford subcortical structural atlases, covering only gray matter tissue (N = 132), containing 8 646 unique functional elements. We filtered the functional elements containing the OFC (either left or right hemispheres), resulting in a set of 261 neuroimaging variables. Individual Fisher’s Z-scores were calculated according to Weissenbacher et al. (48).

### Behavior

We used the Information Sampling Task (IST) to measure impulsive/decision-making behavior (age = 10.32 ± 1.39 years). It consists of a series of trials, each one presenting a 5 x 5 matrix of gray boxes as well as two large colored boxes at the bottom of a computer screen. The child was told that the winning criterium is correctly picking which color is most prevalent under the gray boxes. Touching a gray box reveals the color underneath. No time limit was given. Each trial ends when one of the large colored boxes is chosen, which immediately uncovers all boxes and display a message informing if the child won or lost. We opted for the “decreasing win” condition, where a correct decision is awarded with 100 points, minus 10 points for every box opened. An incorrect decision results in a loss of 100 points. The primary performance score is the probability that the chosen color would be correct, based on the available evidence at the decision time, i.e., dependent on the amount of information sampled (49, 50). The “decreasing score” was also adjusted for discrimination errors, according to Pokhvisneva et al. (51).

### Statistical Analysis

Mediation analysis was first proposed in 1986 (52) and has since been applied in a wide range of scientific fields, such as psychological and biomedical studies (53). Essentially, it decomposes the total effect of the predictor on the outcome in a direct effect and an indirect effect through a mediator variable. We used a single mediator model to investigate the effect of being born SGA on the IST decreasing score, mediated by an OFC-based resting-state functional element. This design requires the estimation of three coefficients using two linear regressions; (i) the effect of the predictor on the mediator (a), and (ii) the effect of the mediator on the outcome when adjusted for the predictor (b) and the effect of the predictor on the outcome adjusted for the mediator (c’). All regressions are adjusted by sex assigned at birth, as well as the age at MRI scan for “a”, and age at IST assessment for “b” and “c’”. The mediation effect is the product of the “a” and “b” coefficients. We selected potential mediators if they were significantly associated with both the predictor and the outcome when adjusted by the predictor. Then, we created a 95% Confidence Interval (CI95) using bootstrapping (10 000 iterations) on the mediation effect, with significance when the CI95 did not include 0. Participants classified as LGA, or with any missing data were excluded from the analysis. Lastly, we removed individuals with influential data points based on the difference in fits (DFFITS) method (54). All analysis were done in RStudio Server v1.4.1717 and R v4.1.1 (55). We used the STROBE cohort checklist when writing our report (56).

## RESULTS

### Sample description

Approximately 13% (n = 82) of the total MAVAN participants attended functional neuroimaging scans. One observation was excluded due to excess head movement. The neuroimaging sample was comparable to the overall cohort (Table 1). 35 more participants were excluded, 5 being LGA individuals, and 30 due to lacking IST measurements. Finally, one individual had an influential measure in the IST score (DFFITS = -1.874) and was removed from the analysis, resulting in 45 individuals (SGA = 31.11%, Figure 1). The final sample was comparable to the overall cohort in gestational age, pregnancy and birth conditions, maternal education, and household income, however it showed lower maternal age at childbirth, as well as the SGA condition being more prevalent (Table 1). Table 1 shows the descriptive statistics comparing the SGA/AGA groups of the resulting participants. There are no group differences regarding gestational age, sex, birth conditions, household income, maternal education, and substance use during pregnancy. The AGA group had a higher incidence of complications during pregnancy and attended the behavioral sessions at slightly younger age than the SGA group. As expected, differences between groups are in birth morphometric variables, with the SGA group showing lower values. There were no differences between the two groups regarding the different IST outcome measures (Table 2).

**Table 1:**
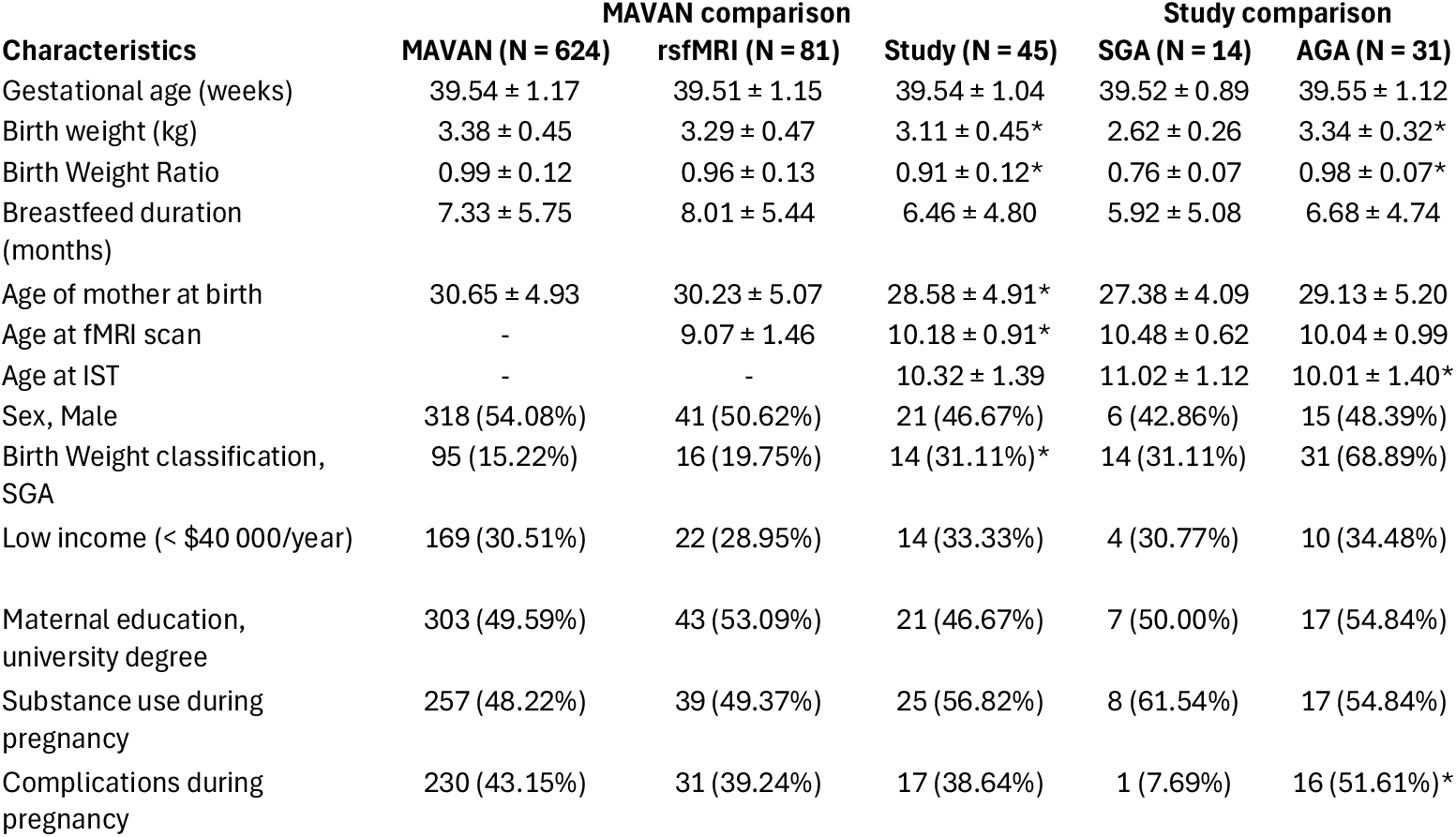
Descriptive statistics. First section compares the neurofunctional (rsfMRI) and study samples with the MAVAN cohort. Second section compares SGA/AGA groups in the study sample. Data presented as “n (%)” or mean ± standard deviation. *statistical significance.

**Table 2:** Information Sampling Task outcome measurements. Data presented as mean ± standard deviation.

| Outcome measurements | Study (n = 45) | SGA (n = 14) | AGA (n = 31) |
| --- | --- | --- | --- |
| Discrimination errors | 1.18 ± 1.37 | 1.12 ± 1.63 | 1.16 ± 1.27 |
| Sampling errors | 2.31 ± 1.29 | 2.00 ± 0.78 | 2.45 ± 1.46 |
| Mean number of boxes opened per trial | 8.28 ± 4.90 | 8.83 ± 5.24 | 8.03 ± 4.81 |
| IST decreasing score | 0.77 ± 0.07 | 0.78 ± 0.08 | 0.77 ± 0.06 |

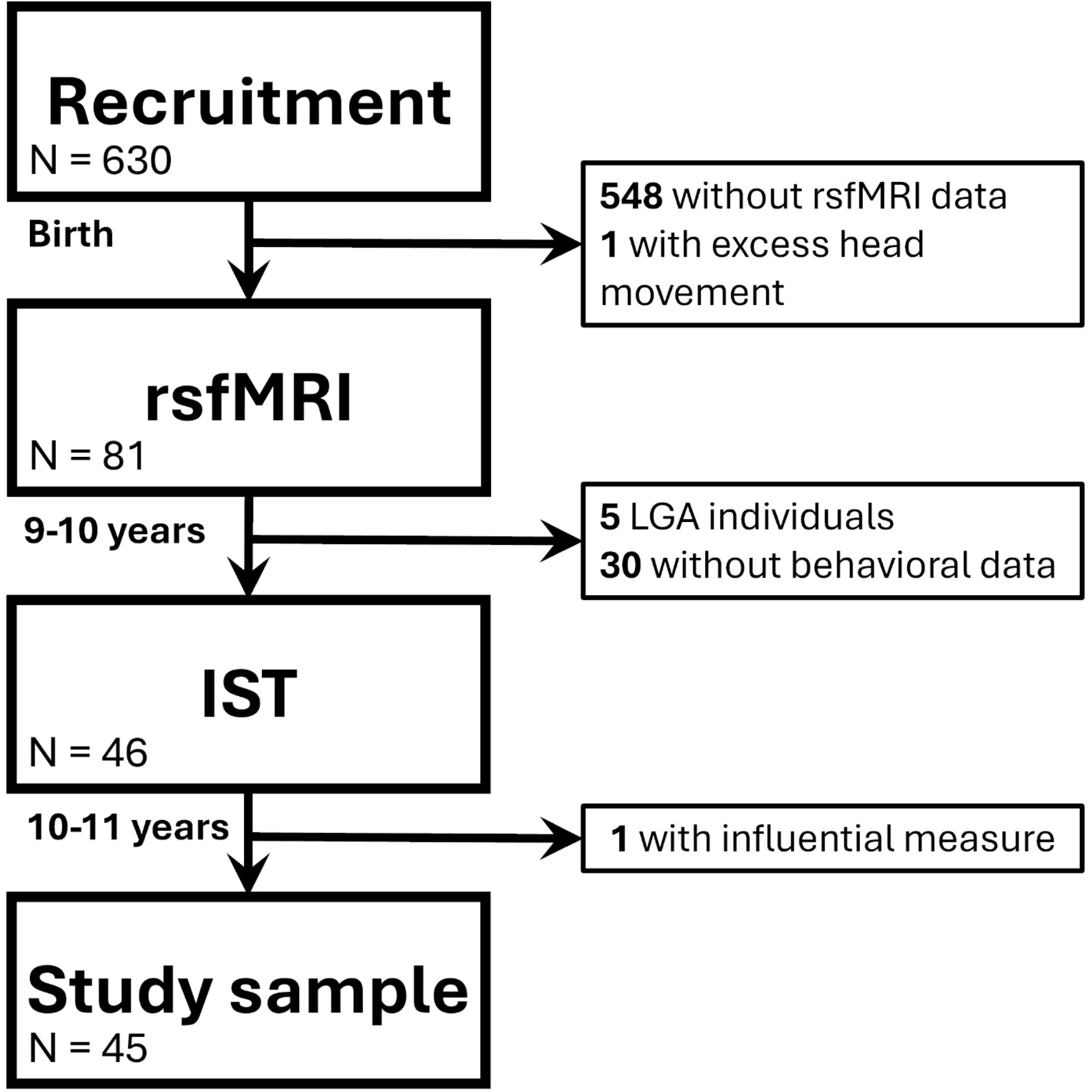

### Mediation analysis

Mediation Analysis decomposes the total effect of a predictor on an outcome in a direct effect, and an indirect effect through a mediator. The design we adopted calculates them via coefficient estimation using two linear regressions: (i) “c’”, effect of the predictor on the outcome adjusted by the mediator; (ii) “a”, effect of the predictor on the mediator, and “b”, effect of the mediator on the outcome conditioned by the predictor. The indirect effect is the product of the “a” and “b” coefficients. Statistical significance is assessed using bootstrapping. We considered only models where both “a” and “b” coefficients were significant, and the confidence interval for the indirect effect did not include zero. We identified a single statistically significant mediation model, with the functional connectivity between the left hemisphere OFC (OFC_L_) and Middle Frontal Gyrus (MFG_L_) mediating the effect of being born SGA on the IST performance (Figure 2). We did not observe a significant direct effect of being born SGA on the IST decreasing score (c’ = 0.043, p = 0.093). The indirect effect through OFC_L_– MFG_L_ was statistically significant (ME = -0.0236 [-0.0556, -0.0022]), suggesting that being born SGA was associated with a decrease in OFC_L_–MFG_L_ functional connectivity (a = -0.139, p = 0.010). At the same time, a higher functional connectivity between OFC_L_–MFG_L_ was linked to a better performance at the task, meaning that children with a higher OFC_L_–MFG_L_ connectivity paced their decision until they gathered enough information to improve their probability of being correct (b = 0.169, p = 0.016). As such, the full model confirms that SGA children display a reduced connectivity between the OFC_L_–MFG_L_ with impact on their decision-making process towards impulsive responses, making the decision when there is less information available, with lower probability of being correct.

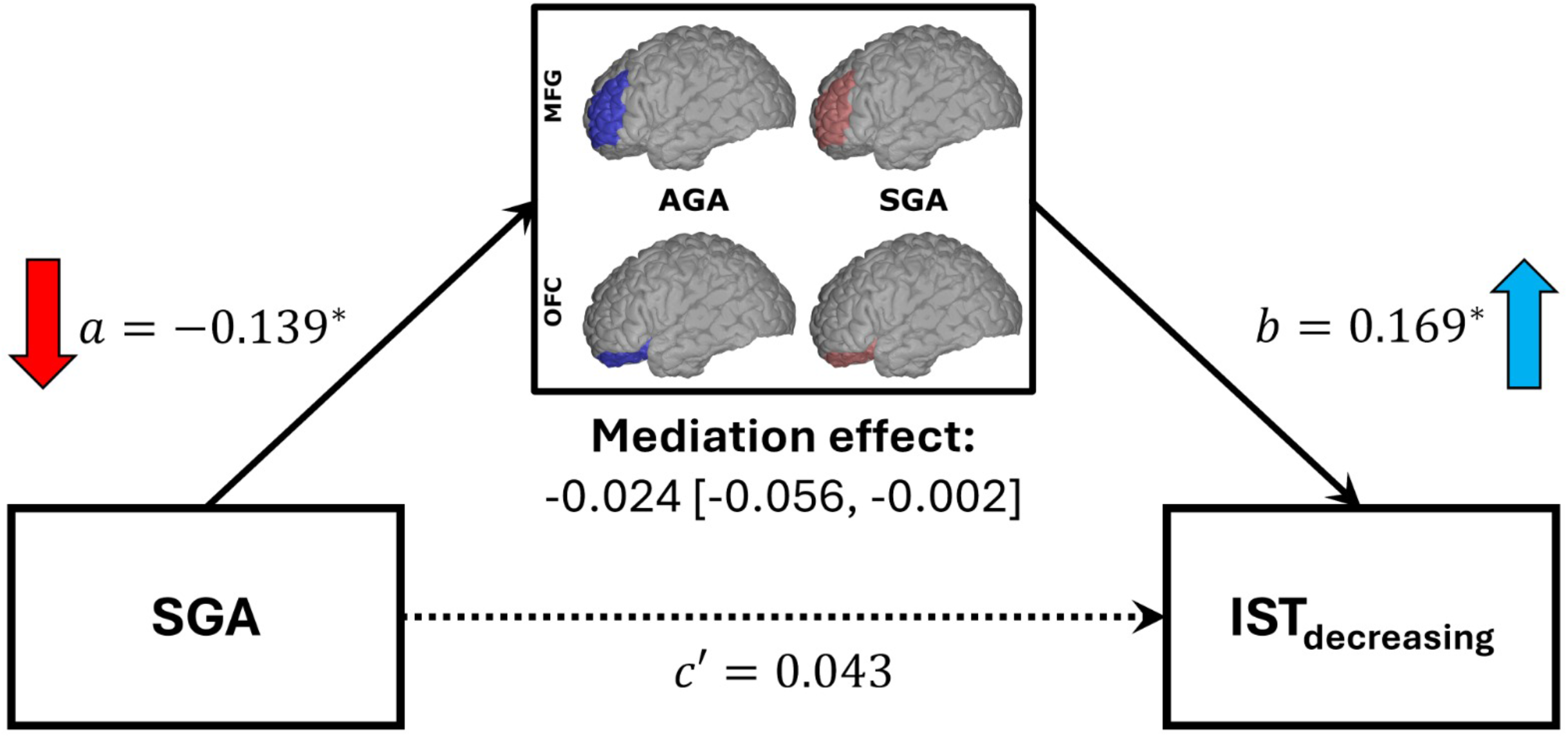

## DISCUSSION

This study is, to the best of our knowledge, the first to investigate the longitudinal association between prenatal adversity, orbito-frontal cortex connectivity and its effects on childhood decision-making processes. The mediation analysis revealed that SGA individuals have altered functional connectivity between the orbito-frontal cortex and the middle frontal gyrus, which results in a more impulsive decision-making behavior. We identified a full mediation framework, i.e., the predictor only influences the outcome through the mediator, highlighting the relevance of the OFC on the effects of poor fetal growth on childhood behavioral outcomes.

The OFC is heavily involved in a wide range of processes related to reward-base decision-making. For instance, it is activated in response to both pleasant and unpleasant rewards stimuli (57), and representation of economic value (58). In addition, it has been suggested that the OFC is involved in learning conditions when there is a delay between the stimulus and the reward (59). Human patients with damaged OFC have increased preference for small-immediate over large-delayed rewards (60), while transcranial stimulation significantly reduces devaluation of delayed rewards (61). Concomitantly, the MFG is linked to attentional processes, with a crucial role in the reorientation of attention and working memory (62). A study investigating individual differences on inhibition of goal-irrelevant information has shown that activation of the left MFG is stronger in the high working memory capacity group, suggesting that it modulates the activation of posterior areas related to sensorial and perceptual information (63). Interestingly, OFC and MFG share the same role on input and modulation of sensorial information to be forwarded to motor areas (20). Thus, it seems that a poorer functional connectivity between the OFC and MFG can be detrimental to attention, with further consequences on downstream processes, e.g., reward-based decision-making. In the current study, this alteration characterizes the group of children born SGA.

It should be noted that SGA is a symptom, not an agent. A lower birth weight respective to the population norm is an indicative of a poor-quality prenatal environment, which has consequences not only at birth, but may manifest later in life, even into adulthood (9). While the multiple factors linked to an SGA makes it a complex concept, it combines a range of neonatal parameters that can be the focus of public health interventions and treatment. If confirmed in future studies, the functional connectivity between the OFC and MFG can be used as a marker for earlier detection of behavioral disorders in the population exposed to prenatal adversity, making it possible to posit individuals at risk on proper treatments.

Early life adversity, such as maltreatment, neglect, or poor fetal growth, has been consistently linked to long-term changes in how people experience rewards and make decisions. These individuals may be more prone to risky decisions and show less flexibility in adjusting their behavior to new information (64, 65). SGA children often prefer immediate, smaller rewards over larger, delayed ones (5, 50), with important consequences for their day to day behaviors (7, 8, 66-68) and long term risk for psychiatric and metabolic disorders (13, 33). Intriguingly, rats exposed to prenatal food restriction — a model for poor fetal growth — also prefer immediate small rewards and show altered dopamine signaling in the OFC, including reduced levels of dopamine D1 receptors (69, 70). This disrupted dopamine function appears to impair how animals learn and respond to value cues (71-76), and functional genomic studies suggest that variations in dopamine signaling affect behavior (77) and risk for metabolic and psychiatric disease (33) in individuals born SGA. Since dopamine plays a central role in motivation and value attribution and processing, these neurobiological changes likely contribute to the behavioral and cognitive effects observed across the lifespan in those affected by early stress.

This work is not free from limitations. Statistical models are always less complex than reality, but they can still be useful (78, 79). Mediation analyses are powerful tools to investigate a chain of events, but even a carefully designed model may fail due to unmeasured confounders. On the other hand, adding too many covariates undermine the model’s statistical power. In this study, descriptive statistics revealed an age difference between SGA/AGA groups at the behavioral assessment, for which we adjusted our models accordingly. In addition, models were adjusted for sex, which is a known factor influencing the effects of SGA on behavior (8, 80, 81). Neuroimaging studies often control for handedness, either as a covariate or excluding left-handed participants (82), but this information is lacking in MAVAN. Another limitation in our study was the sample size. We are aware that our results may suffer from sampling bias, and that a reduced number of observations influences the statistical power and generalizability of our findings. Nevertheless, our study was able to identify a full mediation effect involving the OFC and MFG, influencing the relationship between being born SGA and performance on a computerized task. We strongly endorse replication of this association in larger cohorts. The limited sample impeded an in-depth analysis of sex-specific effects. Lastly, between birth and neuroimaging assessment there is a span of nearly a decade, therefore it may be possible that other environmental effects during childhood are confounding the analysis.

In conclusion, we observed an association between impulsive decision-making and the functional connectivity between the OFC and MFG in response to a poor-quality prenatal environment. Neuroimaging focusing on these areas may identify individuals prone to impairments in their mental health later in life before onset, and direct for proper interventions.

## Supporting information

Figure descriptions

## Data Availability

All data produced in the present work are contained in the manuscript.

## Acknowledgments

This project was supported by the JPB Foundation through a grant to the JPB Research Network on Toxic Stress: A Project of the Centre on the Developing Child at Harvard University. This work was also funded by the Fonds de recherche du Québec – Santé and Canadian Institutes of Health Research (CIHR, PJT-166,066 and PJT-173,237, PI Silveira PP). The authors have no conflicts of interest to disclose.

## Author contributions

**João Paulo M. Hoppe:** Conceptualization, Data curation, Software, Formal analysis, Investigation, Methodology, Visualization, Supervision, Writing – original draft, Writing – review & editing. **Olivia Ruge:** Conceptualization, Investigation, Writing – original draft. **Roberta Dalle Molle:** Conceptualization, Writing – original draft. **Guillaume Elgbeili:** Data curation, Formal analysis, Methodology, Software. **Qizhou Xia:** Data curation, Software. **Patricia P. Silveira:** Conceptualization, Funding acquisition, Project administration, Resources, Supervision, Visualization, Writing – original draft, Writing – review & editing.

## Notes

### Competing Interest Statement

The authors have declared no competing interest.

### Funding Statement

This study was funded by the Fonds de recherche du Quebec - Sante and Canadian Institutes of Health Research (CIHR, PJT-166,066 and PJT-173,237, PI Silveira PP) and supported by by the JPB Foundation through a grant to the JPB Research Network on Toxic Stress: A Project of the Centre on the Developing Child at Harvard University.

### Author Declarations

Research Ethics Board approval was received from the Douglas Mental Health University Institute (IUSMD-03-45, IUSDM-13-09). In the current study, only children from Montreal were included.

