## Supplementary material for "Orbito-frontal cortex functional connectivity mediates the relationship between fetal growth and childhood impulsivity": Figure descriptions

### FIGURE CAPTIONS

**Figure 1:** Sample size flowchart from the MAVAN cohort. IST: Information Sampling Task; rsfMRI: resting-state functional MRI.

**Figure 2:** Mediation model describing that the effect of being born small for gestational age (SGA) on the performance of a decision-making task is mediated by the resting-state functional connectivity between the left orbito-frontal cortex and middle frontal gyrus. Mediation effect is the product of coefficients “a” and “b”, followed by a 95% Confidence Interval.
